# Statistical Methods for Estimating the Protective Effects of Immune Markers Using Test-Negative Designs

**DOI:** 10.1101/2025.04.05.25325304

**Authors:** Casey Middleton, Daniel B. Larremore

## Abstract

While widely used to study vaccine effectiveness, test negative designs (TND) also provide a platform for identifying and quantifying immunological correlates of protection against disease. A key component of such studies is the protection function, the mathematical relationship between the value of an immunological assay and the relative risk of disease. This function is often estimated using logistic regression, comparing the odds of disease at a given assay value to the odds at assay value zero. Here, we show through mathematical analysis and simulation experiments that logistic regression, while common, fundamentally constrains the functional forms of protection that can be inferred from data in TNDs, potentially leading to overly simplistic estimates of the protection function. To address this limitation, we adapt and analyze a scaled logit model, originally developed for case-control data, as a flexible alternative that allows for greater flexibility in estimating protection functions from TND data. We demonstrate that this approach improves accuracy across a range of biologically plausible protection functions, highlight conditions under which it may fail, and provide practical guidance for researchers to adopt it as a new standard for TND studies evaluating correlates of protection.

## Introduction

Antibodies are a key component of the adaptive immune system, equipping the host to quickly recognize and respond to pathogen invasion. Antibodies have been identified as a correlate of protection against infection or severe disease for pathogens such as flu [1], SARS-CoV-2 [2–7], Ebola virus [8, 9], and HIV [10]. While the concentration of antibodies and other immune molecules can be qualitatively linked to decreased risks, establishing the quantitative relationship between immune markers and protection from empirical data is more challenging.

Test-negative design (TND) studies provide cost-effective data to understand the risk reduction provided by potential correlates of protection, and have been widely used to empirically estimate vaccine effectiveness [11–16]. TNDs recruit individuals who seek care for a particular symptom set, such as influenza-like illness (ILI) or fever. All recruited individuals are tested for a pathogen of interest, and the results of the test are reported alongside potential correlates of protection, such as vaccination status. These methods have been extended to understand the association between immunological assays, such as antibody titer, and risk reduction [1–6].

TNDs typically use logistic regression to estimate the association between scalar covariates and risk [1–6]. Logistic regression provides a powerful statistical tool to estimate the odds of a binary outcome, such as observable disease, given covariates such as antibody titer. However, little work has been done to understand the ability of logistic regression to recover widely assumed antibody-protection relationships, whose mathematical forms we refer to as *protection functions*.

In this manuscript, we make progress on the problem by first showing which mathematical relationships— that is, which functional classes of protection functions—may be well suited for inference using logistic regression in TND studies, and which may not. Importantly, while the protective effects of antibodies are typically assumed to be sigmoidal in the literature [17–19], we demonstrate that standard logistic regression applied to TND data inherently limits the ability to accurately recover this functional form of protection. We then extend the scaled logit model introduced by Dunning [20] to TND data, demonstrating its ability to recover a broader class of protection functions from TND studies, with only a small mathematical change. We explore the accuracy of the scaled logit model for inference of various protection functions, demonstrate its utility on empirical TND data, identify conditions under which it may fail, and provide practical guidance for researchers to adopt this model as a new standard for estimating the protective effects of immunological assays in TND studies.

## Results

### Standard logistic regression recovers only exponential protection functions from TND data

Test-negative designs (TNDs) are commonly used to estimate vaccine effectiveness (VE), defined as the reduction in risk of disease due to vaccination. In TNDs, VE is estimated via the odds ratio (OR) of disease among vaccinated versus unvaccinated individuals, i.e. *V E* = 1 − *OR* [11, 12]. Logistic regression is a convenient statistical method for estimating this odds ratio and is often employed in the analysis of TND data [11, 13–15, 21], primarily because it enables one to include and control for other relevant covariates *θ*. In a logistic regression model, the log-odds of disease given vaccination status and other covariates is modeled as,

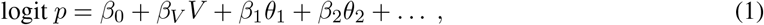

where *p* is the probability of disease, *V* is an indicator for vaccination status, and *θ*_*i*_ represent other covariates. The coefficient *β*_*V*_ represents the change in log-odds of disease associated with vaccination, meaning the odds ratio is 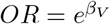. Therefore, vaccine effectiveness is defined as,

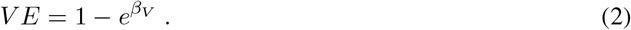

When analyzing scalar correlates of protection, such as antibody titers, an intuitive substitution is to simply replace the binary vaccination status *V* with a measured titer *A* and proceed with typical logistic regression [1–6],

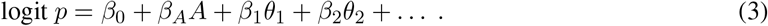

Relative to a baseline risk when *A* = 0, this replacement yields a protection function Φ, which describes the fraction by which risk is reduced as a function of *A*,

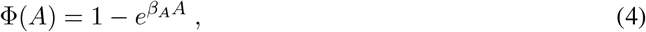

where *β*_*A*_ is the change in log-odds of risk given a one unit increase in antibody titer.

Equation (4) provides the exact functional form of the induced protection function when logistic regression is used to learn from TND data. Consequently, logistic regression is well suited to recovering this particular functional relationship between titers *A* and protection Φ. Hereafter, we refer to (4) as an exponential protection function.

To illustrate how logistic regression accurately recovers exponential protection functions, we developed a straightforward synthetic TND data testbed. First, we specify the shape of a protection function, and here we assume the exponential form (Eq. (4); Fig. 1A) with some *β*_*A*_ < 0. Second, we use this protection function to simulate infected and uninfected antibody titer distributions (Fig. 1B). In this simulation, we assume a large sample size (*N* = 10^5^) and perfect diagnostic sensitivity and specificity when assigning infected status. Third, we use logistic regression to estimate infection risk (Eq. (3)) from the simulated data (Fig. 1C). Lastly, we use this risk estimate to compute the odds ratio with a baseline of *A* = 0 in order to arrive at an estimated protection function (Fig. 1D). This simulation illustrates what we have shown mathematically: using logistic regression to infer risk can recover the true protection function when that function is exponential. Statistically speaking, when the logistic regression model is correctly specified for the true protection function, estimation is consistent.

**Figure 1.**
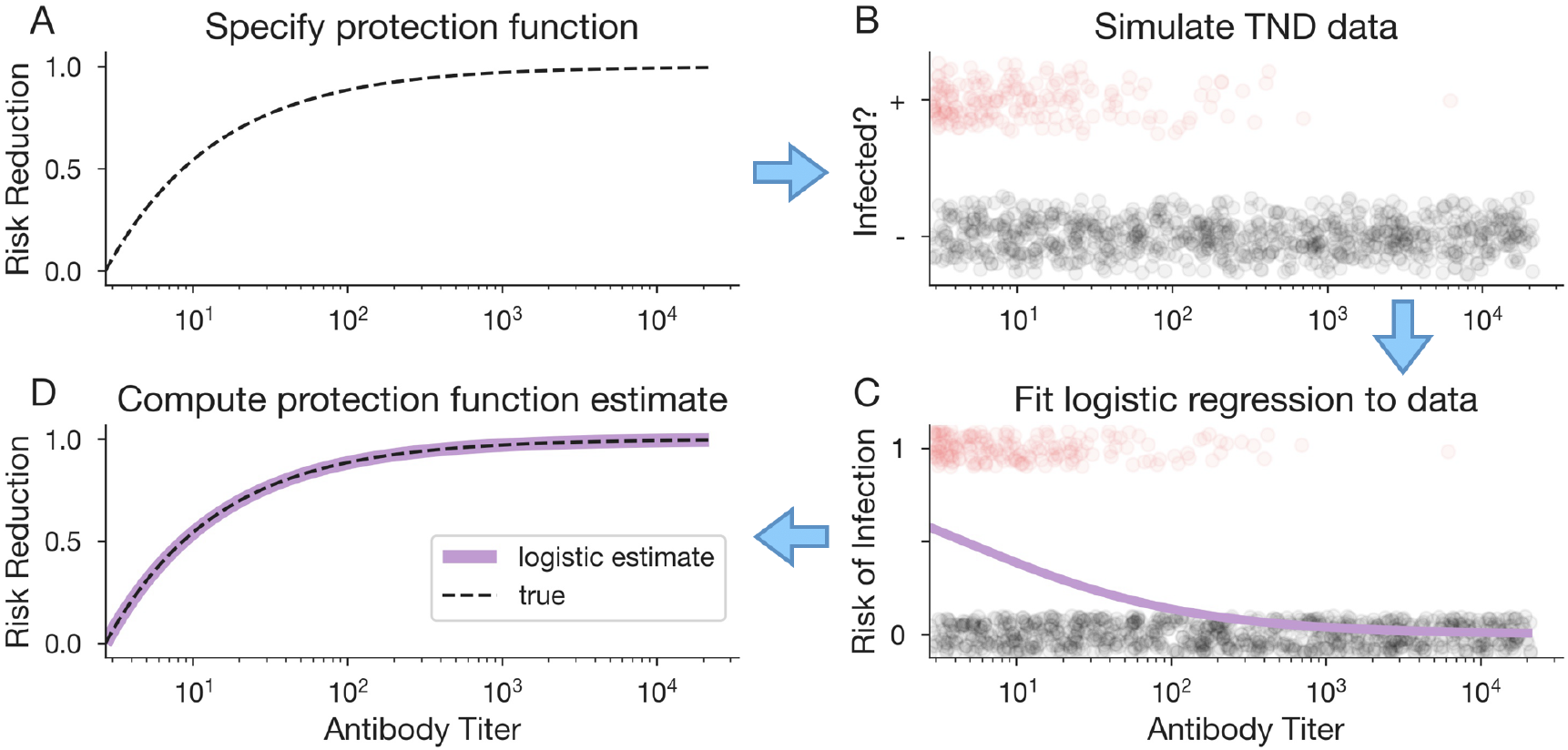
Logistic regression accurately estimates exponential protection functions. (A) First, we specify an exponential protection function using Eq. (4) and some *β*_*A*_ < 0. (B) Next, we simulate TND data using this protection function to produce antibody distributions for test-positive (red circles) and test-negative (gray circles) individuals (subset of data shown here). (C) Logistic regression is used to estimate risk of infection from the simulated data via Eq. (3). (D) Lastly, we compute the estimated protection function from the logistic regression model results (purple curve), which we compare to the true protection function (black dashed curve) that was used to generate the data, specified in step A.

### Logistic regression is poorly suited to recover a sigmoidal protection function from TNDs

The fact that logistic regression induces to a protection function defined by a single-parameter exponential (Eq. (4)) raises an important question: what if the true protection function is not exponential? We illustrate this type of mismatch between model and data—a model misspecification problem—through a second simulation experiment which follows the same four step procedure illustrated above. However, in this experiment, we generate data according to a plausible (but non-exponential) alternative protection function, and then fit a logistic regression model to the data.

Rather than an exponential protection function, we used a sigmoidal shape (Fig. 2A), as is commonly assumed in the literature [17–19]. The sigmoidal protection function is defined by

**Figure 2.**
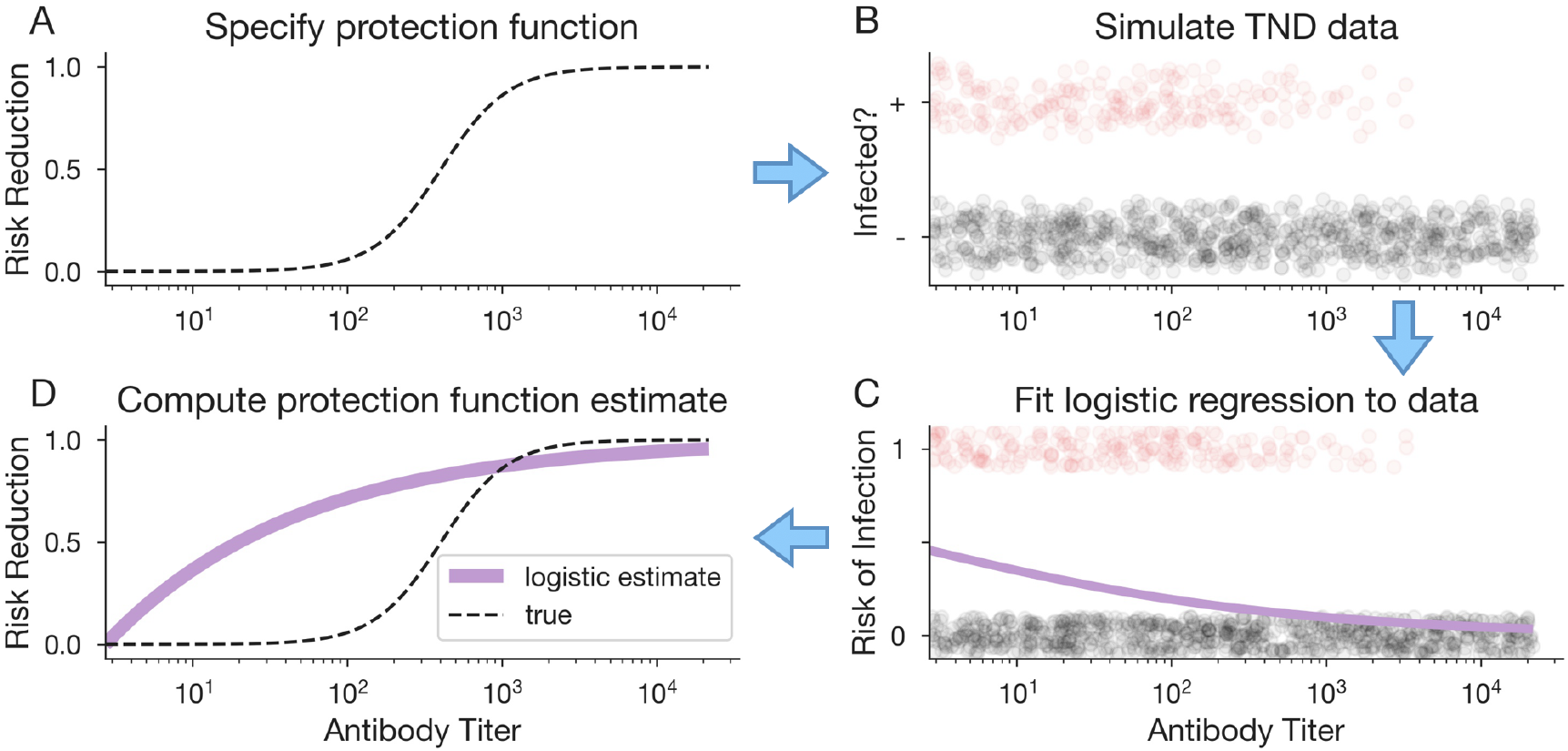
Logistic regression does not accurately estimate sigmoidal protection from TND data. (A) First, we specify a sigmoidal protection function using Eq. (5) and some *β*_0_ > 0, *β*_*A*_ < 0. (B) Next, we simulate TND data using this protection function to produce antibody distributions for test-positive (red circles) and test-negative (gray circles) individuals (subset of data shown here). (C) Logistic regression is used to estimate risk of infection from the simulated data via Eq. (3). (D) Lastly, we compute the estimated protection function from the logistic regression model results (purple curve), which we compare to the true protection function (black dashed curve) that was used to generate the data, specified in step A.

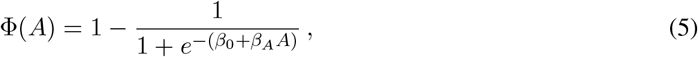

for some *β*_0_ > 0, *β*_*A*_ < 0. We simulated TND data using this sigmoidal protection function (Fig. 2B), and fit a logistic regression model to the synthetic data (Fig. 2C). The resulting estimated protection function (Fig. 2D) is no longer accurate.

Logistic regression fails to recover the true sigmoidal protection function used to generate the data, as evidenced by the mismatch between the estimated and true curves (Fig. 2D). These results are not unexpected: the logistic regression model is restricted to exponential protection functions (Eq. (4)) and thus cannot recover other functional forms, even when applied to an ideal dataset with large sample size and no measurement error. To infer a potentially non-exponential relationship between antibodies and protection, a different statistical approach is required.

### Alternative models for correlates of protection inference

Due to the inflexibility of standard logistic regression, a few alternatives have been proposed for inferring the functional relationships for correlates of protection, which we briefly review. Zhang et al. show that rescaling model inputs such that logit *p ~ ln*(1 − *X*^*n*^) for rescaled titers *X ∈* [0, 1] can recover a linear protection function from TND data when *n* = 1, and matching the power *n* to the protection function can recover polynomial functions [22]. This change increases model flexibility to capture polynomial protection functions, given an a priori choice of *n*. Alternatively, the authors propose the use of semi-parametric generalized additive models (GAMs) as a highly flexible inference approach.

One might also consider using higher order polynomial terms in a typical logistic regression model, such that,

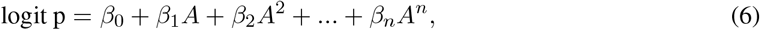

an approach commonly employed when the relationship between the predictor and log-odds is nonlinear. This approach also increases model flexibility, but it has the drawback of overfitting data for large *n* and requires model selection to determine the appropriate number of polynomial terms.

For case-control data, Dunning proposed a scaled logit model [20] which defines the probability of observing a case as

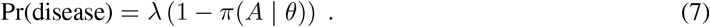

Here, *λ* denotes the probability a fully susceptible individual develops disease, and *π*(*A* | *θ*) describes the probability that an individual with titer *A* is protected, given model parameters *θ*. If a logistic protection function is assumed for *π*(*A*), this results in risk of disease,

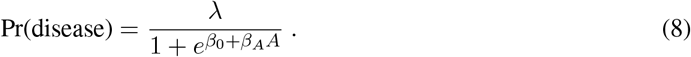

Thus, the scaled logit model has a similar shape to the logistic, but the parameter *λ* reduces the expected force of exposure for individuals with low antibody titer.

### The scaled logit model can recover many functional relationships between antibody titers and risk reduction from TND data

We now extend the scaled logit model for use in TNDs by substituting the risk function in Eq. (8) into the protection function estimator, i.e. one minus the odds ratio of protection at titer *A* vs titer *A* = 0. This yields an estimated protection function of

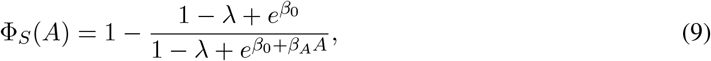

where subscript *S* identifies this as the estimated protection function using the scaled logit model. Importantly, the scaled logit requires estimation of only one more parameter *λ* than typical logistic regression, and this, in turn, produces a protection function with three parameters *β*_0_, *β*_*A*_, and *λ* [Eq. (9)] instead of only one parameter *β*_*A*_ given by logistic regression [Eq. (4)]. Consequently, we may expect protection functions resulting from the scaled logit model to be substantially more flexible in modeling correlates of protection.

To demonstrate the flexibility of a scaled logit for TND studies, we simulated TND data under both exponential and sigmoidal protection functions (Fig. 3A,B) and used the scaled logit model to estimate the protection function for each synthetic data set. We observe that the scaled logit model accurately fits both exponential and sigmoidal protection functions. These results demonstrate that the scaled logit model can recover not only exponential protection, which logistic regression can recover, but also sigmoidal protection functions, which logistic regression cannot.

**Figure 3.**
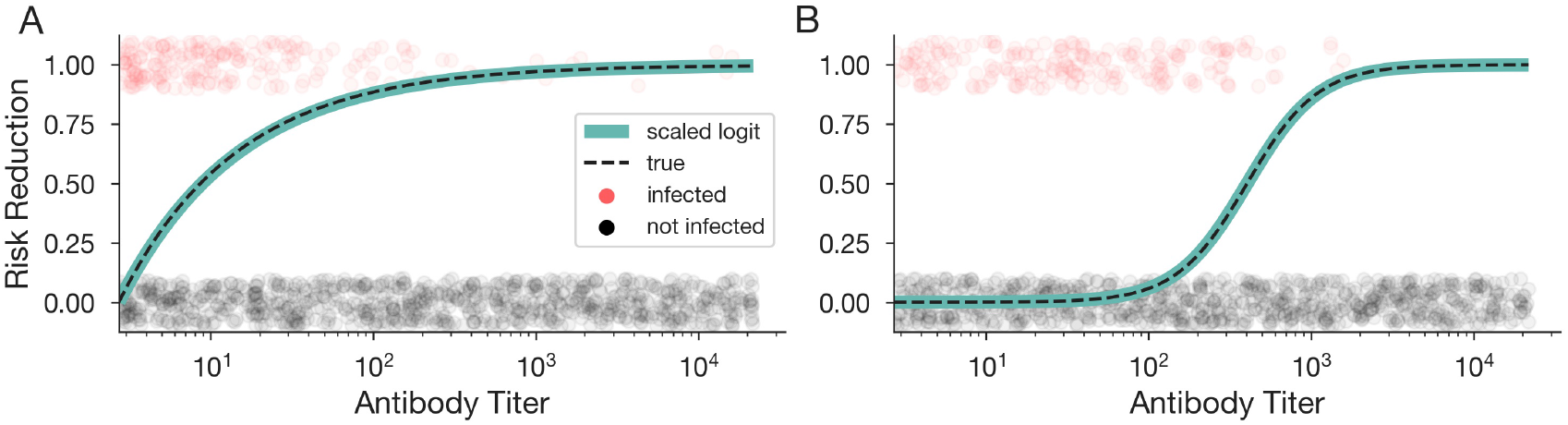
The scaled logit model can recover general antibody protection functions from TND data. Estimated protection function using the scaled logit model (green curve) trained on data generated from exponential (A) and sigmoidal (B) protection functions (dashed black curve). Circles show a subset of infected (red) and uninfected (gray) simulated individuals at various antibody titers used to train the model.

To further explore the ability of these models to accurately recover a variety of protection function scenarios, we subjected both the scaled logit and logistic models to additional synthetic data tests generated from two additional protection functions, a shifted step function and a constant Φ(*A*) = 0 null result test. As with the sigmoidal protection function, the scaled logit model accurately estimated step-function protection, while logistic regression did not (Fig. S1A). Both models accurately fit the null protection function Φ(*A*) = 0, confirming their ability to correctly find a true null result (Fig. S1D). In further tests, we restricted the range of antibody titers such that few individuals had high or low titers, to challenge the scaled logit model’s ability to recover protection functions from less optimal data, but its high accuracy was unaffected (Fig. S2).

These results demonstrate that introducing a single additional parameter in a scaled logit model enables inference of a broader range of functional forms from TND data, including sigmoidal risk reduction, which standard logistic regression cannot capture. Adopting this scaled logit model when estimating the protective effects of immunological assays from TND data would allow for inference of a markedly broader set of possible protection functions. Establishing that the scaled logit model can accurately recover either sigmoidal or exponential protection functions from synthetic data indicates that we may be able to recover either functional form from real data, which we now turn to.

### The scaled logit model estimates sigmoidal curves from empirical TND data

Our synthetic data experiments demonstrate that the scaled logit model can recover a sigmoidal protection curve in TND studies, while logistic regression cannot. To understand how these findings extend to empirical data, we applied the same inference pipeline to published TND data which associates anti-spike antibody titers with SARS-CoV-2 infection, as previously described [23]. This study enrolled 2300 patients between March 22, 2021, and Aug 17, 2022 who presented with undifferentiated acute febrile syndromes across two hospitals in the Dominican Republic. Nasopharyngeal swabs and sera from each patient were tested for SARS-CoV-2 infection using real-time PCR (rtPCR) and for total anti-spike antibodies with the Elecsys platform (Roche Diagnostics, Indianapolis, IN, USA). Positive samples with cycle threshold values less than 25 were sequenced using Oxford Nanopore or Illumina platforms (N=216). These variant annotations allow inference of variant-specific protection functions by comparing variant-positive cases to a random sample of negative controls.

We first estimate five variant-specific protection functions using the scaled logit model stratified by variant (Fig. 4A). Our results show that variants which emerged earlier, i.e. delta and mu, are characterized by protection functions which increase rapidly to provide high levels of protection even at low titers. We estimate 50% risk reduction when anti-spike antibody titers reach 10^1.51^ and 10^1.00^ binding antibody units (BAU)/mL for delta and mu, respectively. The estimated protection functions for the omicron sublineages BA.1, BA.2, and BA.4/5 are more sigmoidal in shape, estimating 50% risk reduction at titer values of 10^3.48^, 10^3.10^, and 10^3.14^ BAU/mL, respectively. We note that stratifying by variant results in relatively small sample sizes and few positive cases, especially for BA.2 and BA.4/5 (17 and 19 cases, respectively).

**Figure 4.**
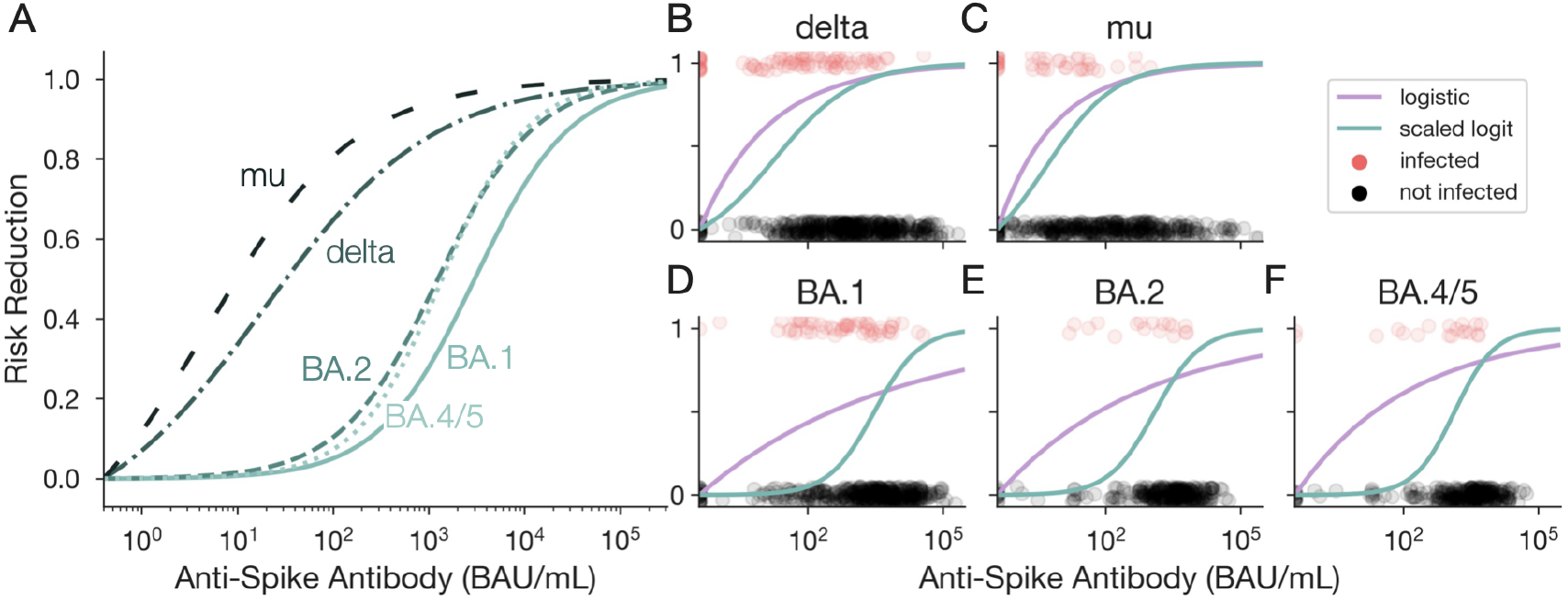
The scaled logit model allows for inference of a sigmoidal protection function from empirical TND data. Estimated protection function using the scaled logit model (green curves) and logistic regression (purple curves) fit to empirical TND data for SARS-CoV-2, stratified by variant of concern for (B) delta, (C) mu, (D) BA.1, (E) BA.2, and (F) BA.4/5. Circles show infected (red) and uninfected (gray) sampled individuals for each variant. BAU, binding antibody units.

Next, we compare the protection function estimated by the scaled logit model to that estimated by typical logistic regression, stratified by variant (Fig. 4B-F). For all variants, we observe that logistic regression produces an exponential protection curve which under-estimates the titer required to achieve 50% risk reduction when compared to the more sigmoidal scaled logit model estimates. This underestimation is substantial for the omicron sub-lineages (BA.1, BA.2, BA.4/5), whose sigmoidal scaled logit protection curves deviate more drastically from the logistic estimates than the delta and mu variants. Given that the scaled logit model can reliably estimate either exponential or sigmoidal protection functions (Fig. 3), the sigmoidal estimates observed across SARS-CoV-2 variants indicate that the true underlying protection mechanisms likely more closely resemble sigmoidal than exponential curves. These empirical results emphasize what we previously showed with synthetic data: the scaled logit model allows for characterization of a broader range of protection functions than logistic regression.

### Scaled logit model accuracy depends on sample size

Studies vary widely in the total number of individuals sampled, as well as the proportion of samples that are cases versus controls [1–6]. Factors such as study duration, disease prevalence, and specificity of the symptoms which trigger enrollment into the study may impact sample size and case-to-control ratios. To explore how these sample attributes impact the accuracy with which we can infer a protection function, we simulated TND data from a range of sample sizes and case-to-control ratios using a sigmoidal protection function, and the simulated data was used to estimate the protection function from both the scaled logit and logistic regression models. For each scenario, we computed error as the discrete *ℓ*_2_ norm of the difference between the best-fit model 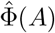 and the true protection function Φ_true_(*A*), given by

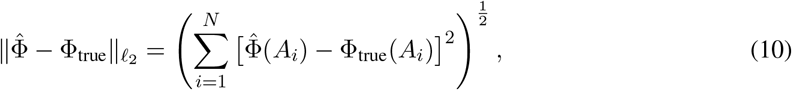

where 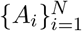 is the set of all observed antibody titers in the study. To account for stochasticity in the generation of synthetic data, the mean error was calculated for 50 simulations per scenario.

When the scaled logit model is used to estimate protection, we observe higher error for smaller sample sizes (Fig. 5A) due to model overfitting. At low sample sizes, the model is unable to capture the true slope of the sigmoid (panel I). As sample size increases, model fits more closely resemble the true protection function on average (panels II, III). Furthermore, we observe lower error when a higher proportion of total samples are cases (lighter colored curves), especially at low sample sizes, especially at sample sizes below 1000 (Figs. S3, S4). This demonstrates that cases typically provide more information to model inference than controls.

**Figure 5.**
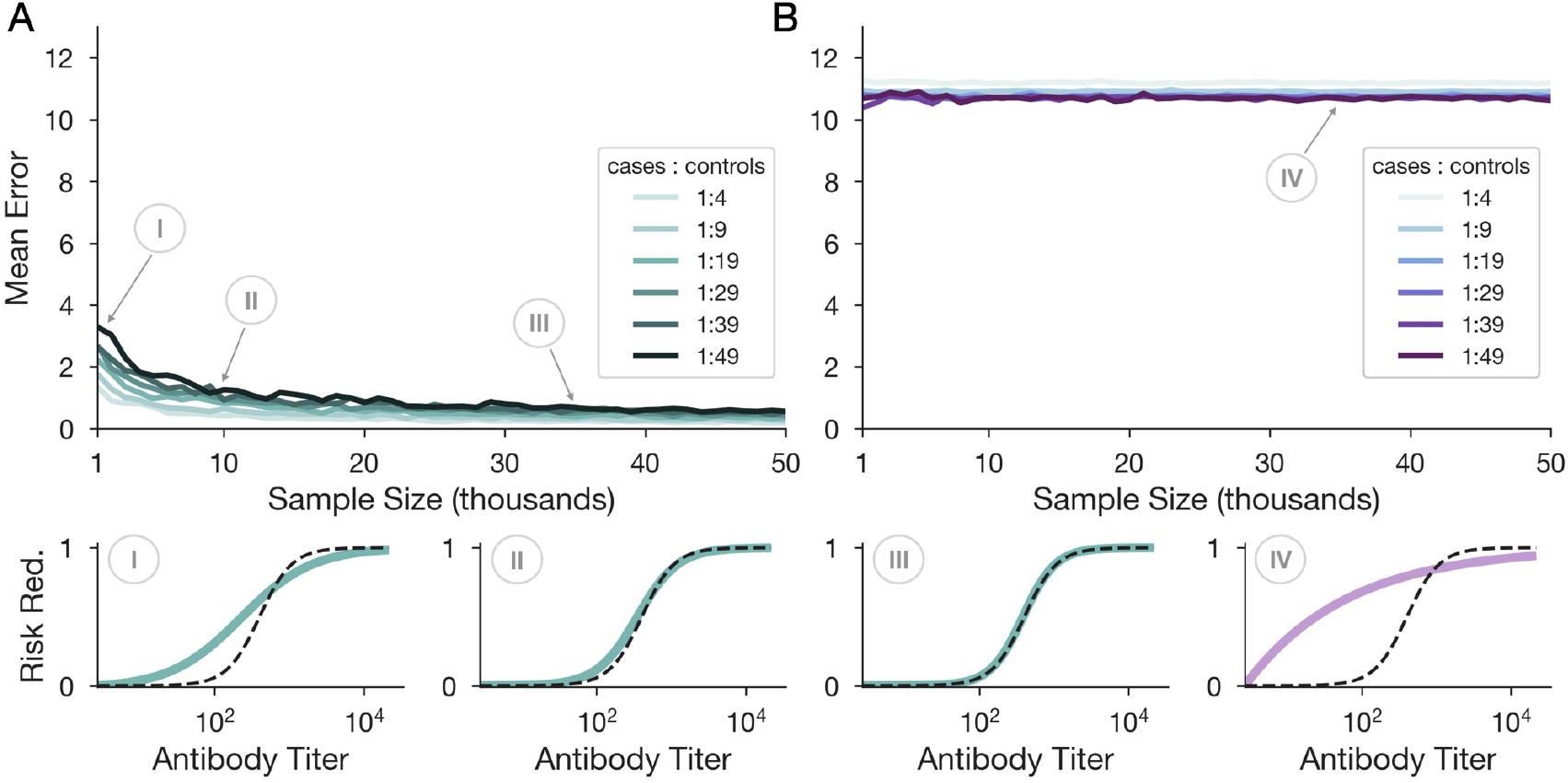
The scaled logit model is better specified to recover sigmoidal protection from TND data than logistic regression, irrespective of sample size. Mean error over 50 simulations per scenario for various sample sizes and case-to-control ratios using the scaled logit model (A) and logistic regression (B). Error was defined as the discrete *ℓ*_2_ norm (Euclidean distance) between the modeled and true protection functions. Darker colored lines represent scenarios with more controls per case. Insets show protection function estimates (colored curve) compared with the true protection function (black dashed curve) from a single simulation when *N* = 1, 10, and 35 thousand for the scaled logit (I-III) and 35 thousand for the logistic model (IV) using a 1 : 49 case-to-control ratio.

The same analysis of error in protection function estimation using logistic regression shows uniformly high error across all scenarios (Fig. 5B), which does not approach 0 as sample size increases. These results reflect the large discrepancy between the inferred and true protection functions when using logistic regression to estimate a sigmoidal protection function from TND data (panel IV). This observation offers further support that logistic regression is poorly suited to recover a sigmoidal protection function, at any sample size, due to model misspecification.

### The scaled logit model has poor accuracy if antibodies do not provide perfect protection

While the addition of a scaling constant *λ* allows for more flexible protection function estimates, Φ_*S*_ still approaches 1 as *A → ∞* (Eq. (9)) for typical monotonically decreasing risk parameterization (i.e. *β*_0_ < 0, *β*_*A*_ > 0). Because of this, the model may be ill suited to recover scenarios where protection is imperfect, or where near-perfect protection exists but is simply not represented in the sampled controls.

We conducted two numerical experiments, each of which explores a different scenario in which the scaled logit may have poor accuracy. In the first experiment, we asked what happens if the protection function does not saturate to one — that is, to perfect protection — as antibody titers grow large. We simulated TND data under a sigmoidal protection function which saturates at 65% (Fig. 6A). We then estimated the protection function using the scaled logit model. The best-fit protection function overestimates protection at both low and high antibody titer, and approaches one as titer values increase. Varying the saturation point, or maximum possible protection, we observe relatively stable error estimates until the maximum possible protection surpasses 90%, at which point error approaches 0 (Fig. 6B).

**Figure 6.**
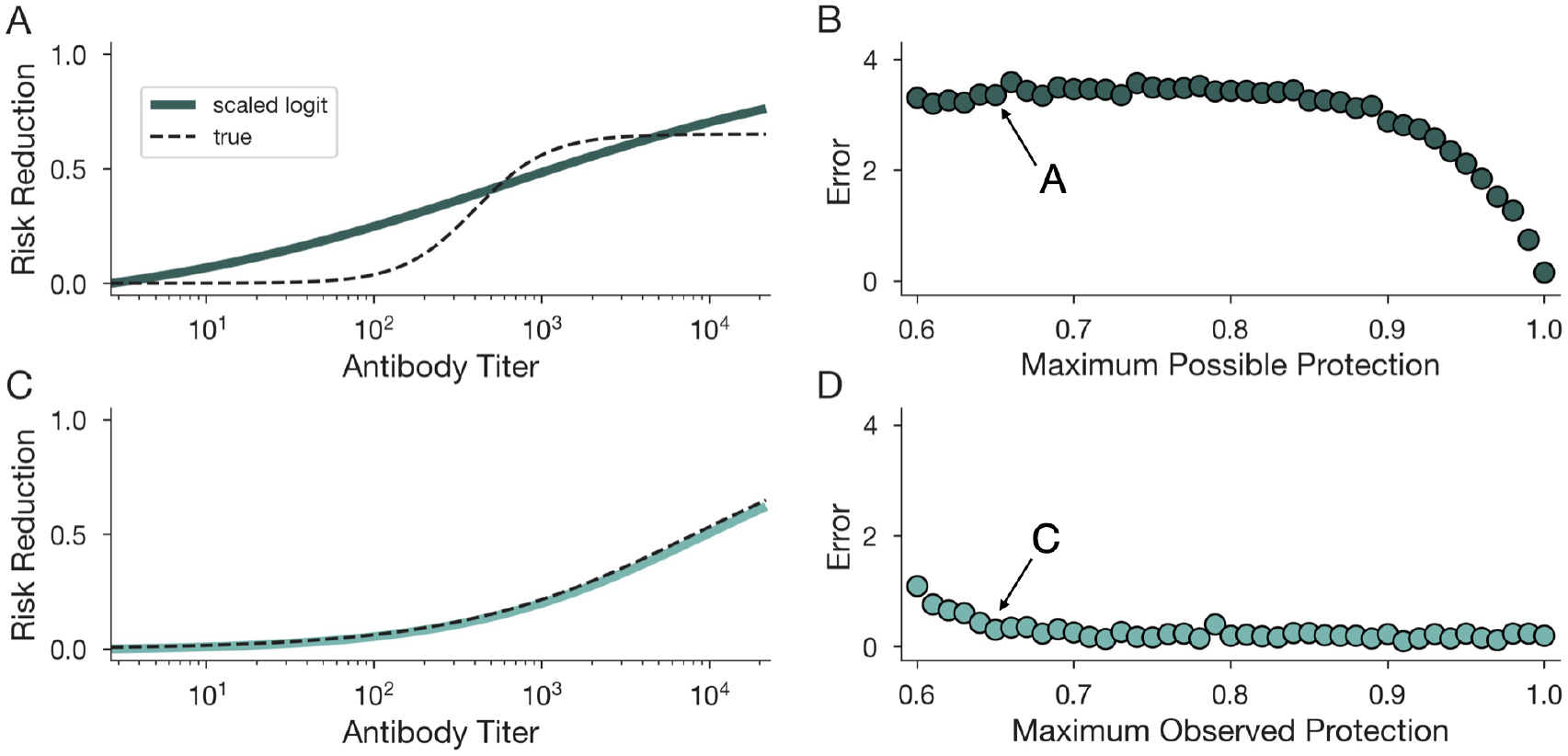
Scaled logit model has reduced accuracy if antibodies do not provide perfect protection. The scaled logit model is fit to TND data generated from a sigmoidal protection function which does not saturate to 1 (A,B) and when individuals with near-perfectly protective antibody titers are not sampled among controls (C,D). (A,C) Predicted protection functions from the scaled logit model (green curves) versus true protection (black dashed curves) for a maximum protection of 0.65. (B,D) Prediction error (Euclidean distance between predicted and true protection) under varying maximum protection levels.

In the second experiment, we asked what happens if individuals with near-perfectly protective antibody titers are simply not sampled among the controls. To simulate this, we generated TND data where the maximum sampled antibody titer is *A*_max_, but the slope of the sigmoidal protection function is reduced such that risk reduction is perfect only for values *A ≫ A*_max_ (Fig. 6C). For this scenario, we observe an estimated protection function that nearly perfectly estimates the true protection function. Varying the maximum observed protection shows that error is low and nearly constant across scenarios (Fig. 6D). Comparing across experiments, we observe higher error for saturating protection (Fig. 6B) than for unsampled perfect protection (Fig. 6D). Distinguishing between these two scenarios may facilitate a better understanding of confidence in model fits.

## Discussion

Our results establish the scaled logit model as a more flexible alternative to typical logistic regression methods for estimating protection as a function of scalar immunological assays in test-negative design (TND) studies. Using simulated data, we see that the scaled logit model can recover not only an exponential protection function, which logistic regression can recover, but also a sigmoidal protection function, which logistic regression fails to recover. These findings extend to real data, where we observe sigmoidal estimates of the protection function when characterized using the scaled logit model, compared to exponential estimates using logistic regression. These results lead us to recommend the adoption of the scaled logit model for modeling scalar correlates of protection from TND data.

Despite its increased flexibility, we also demonstrate that the scaled logit model may not accurately recover the true protection function for sample sizes below 1000, and that model consistency deteriorates for sample sizes under 10^4^. Furthermore, we show that if protection does not saturate to 1, model accuracy may suffer. Alternative sigmoidal models with greater flexibility to capture protection functions which do not saturate to 1, exhibit asymmetry, or are characterized by extremely steep transitions have been previously explored [24], but may require model selection or prior knowledge of the expected shape of the protection curve. Alternative innovations using generalized additive models (GAMs) have also been proposed with promising results [22].

Our methods are subject to a number of limitations. First, we simulate data under ideal sampling scenarios, exploring limitations of sample sizes and case-to-control ratio variations, but not sensitivity and specificity of diagnostic tests [15, 25] or noise in immunological assay measurements. Second, we do not consider the effects of demographic covariates, such as age and sex, or immunity covariates, such as time since last infection or other immunological assays [26]. While these covariates may impact protection estimates, we posit that the scaled logit model can be used to control for their effects by including additional terms in Eq. (8). Third, our exploration of protection functions is limited to functions which start at 0 when *A* = 0 and monotonically increase with increasing titer. These functions do not account for immune imprinting [27] or antibody-dependent enhancement [28], which may lead to non-increasing protection functions. Lastly, our synthetic data testbed does not account for differences in care-seeking behavior or potential cross-protection between circulating pathogens, and assumes that protection is “leaky” instead of “all-or-nothing,” which can lead to differential depletion of susceptibles over the course of a season. These factors have been shown to potentially bias the protection estimates inferred using test-negative designs for vaccine effectiveness studies [11–13, 29] and would likely also bias study results inferring correlates of protection.

Quantifying the relationship between assay value and risk reduction is an important task to understand the utility of immunological assays. Informative immunological assays may open up new data streams to model heterogeneous susceptibility in immune-experienced populations [30,31] and vaccination strategies [32,33]. Furthermore, understanding the strength of correlations between titer values and protection could be used to predict vaccine efficacy from titers alone instead of following a cohort in vaccine trials, reducing costs and increasing the speed at which trials may be performed. The scaled logit model, in combination with test-negative designs, may provide more accurate mathematical descriptions of how immunological measurements predict protection levels, opening the door for immunological assays to inform infectious disease modeling efforts, and establishing quantitative measurements of correlates of protection as substantially higher in value than binary alternatives [34].

## Materials and Methods

As described in the main text, our simulation test bed follows four key steps: (i) specify the shape of the protection function Φ(*A*); (ii) simulate antibody titers for infected and uninfected subjects of a test-negative design study; (iii) estimate a model, either logistic regression or the scaled logit; and (iv) use the estimated model to compute a protection function. These materials and methods provide additional details and code for these steps.

### Protection functions

We considered four protection functions. These are the exponential, 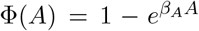; the sigmoidal,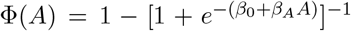; the step-function or threshold, Φ(*A*) = Θ(*A* − *A*_0_), where Θ is the Heaviside function; and the constant null model, Φ(*A*) = 0.

### Test-negative design data simulations

TND studies typically recruit all individuals who report a particular symptom or set of symptoms. Recruited individuals are tested for a pathogen of interest, with data reported on: (1) test result for pathogen of interest (positive or negative), (2) value of covariate(s) assumed to be associated with infection risk (i.e. vaccination status or immunological assay), and (3) other variables which may impact risk (i.e. age, sex). Our simulations focus only on quantitative antibody titer *A* as the covariate of interest and ignore other covariates.

To generate titer distributions for test-negative controls, we drew *A* from a known distribution. In the main text, ~ *log*(*A*) ~ log-uniform(1, 10), and in supplemental analyses, *A* ~ uniform(1, *e*^10^) and *A* ~ lognormal(5.5, 2). To generate titer distributions for test-positive cases, we used a rejection sampling technique such that a candidate titer was drawn from the same distribution as controls, and was then rejected with probability 1 − Φ(*A*), simulating the protective effects of those antibodies. Because cases and controls can be generated by these procedures independently, samples were drawn in order to achieve a specified total sample size *N* as well as a chosen case-to-control ratio *c*.

### Fitting logistic regression models

The logistic model in Eq. (3) was fit to simulated TND data using `sklearn` (Python v3.8.11) without regularization (see Code Availability). Parameters were fit to log titer data, following typical procedure [1, 4–6]. The protection function was then computed using the odds ratio definition 1 − OR, comparing the odds of testing positive with antibody titer *A* against the baseline odds of testing positive at *A* = 0.

### Fitting scaled logit models

The scaled logit model in Eq. (8) was fit to simulated TND data using maximum likelihood estimation (MLE). The likelihood of parameters given TND data is,

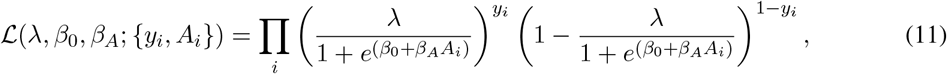

where *A*_*i*_ denotes individual antibody titers and *y*_*i*_ *∈* {0, 1} denotes an observed non-infection or infection, respectively. We wish to find the parameters (*λ, β*_0_, *β*_1_) which maximize this likelihood function, or equivalently the parameters that minimize the negative log-likelihood,

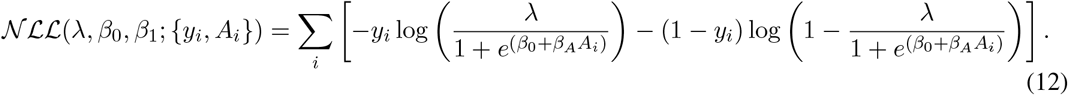

This negative log likeilhood was fit to TND data using `scipy.minimize` (Python v3.8.1), implemented with the Nelder-Mead minimization algorithm [35] and options to increase the number of iterations before convergence. To prevent overfitting of small sample sizes, which results in a large *β*_*A*_ slope parameter, we include regularization on the term 0.1 × *β*_*A*_ ^2^. Both Python and R implementations are provided in Supplementary Materials. The protection function was then computed using the odds ratio definition 1 − OR, comparing the odds of testing positive with antibody titer *A* against the baseline odds of testing positive at *A* = 0.

## Data Availability

All synthetic data in the present study can be reproduced using the code available at https://github.com/CaseyMiddleton/TNDforCOP.

## Code and data availability

Implementations of scaled logit model fitting in R and Python are provided in Supplementary Materials. All code used for data simulation, model fitting, and reproducibility, as well as empirical data used in the study, can also be found at github.com/CaseyMiddleton/TNDforCOP.

## Acknowledgments

C.E.M. was supported in part by the Interdisciplinary Quantitative Biology (IQBio) program at the University of Colorado Boulder. D.B.L. was supported in part by an NSF Alan T. Waterman Award (SMA-2226343) and the US Centers for Disease Control and Prevention (NU38FT000008). The funders played no role in the design, conduct, or reporting of this study. We thank Drs. Sarah Cobey, Scott Olesen, and Edward Schrom for their feedback on earlier versions of this work, and Dr. Eric Nilles for making empirical data publicly available for re-analysis in our manuscript.

## Ethics declaration

The authors declare no conflicts of interest.

## Supplement

**Figure S1:**
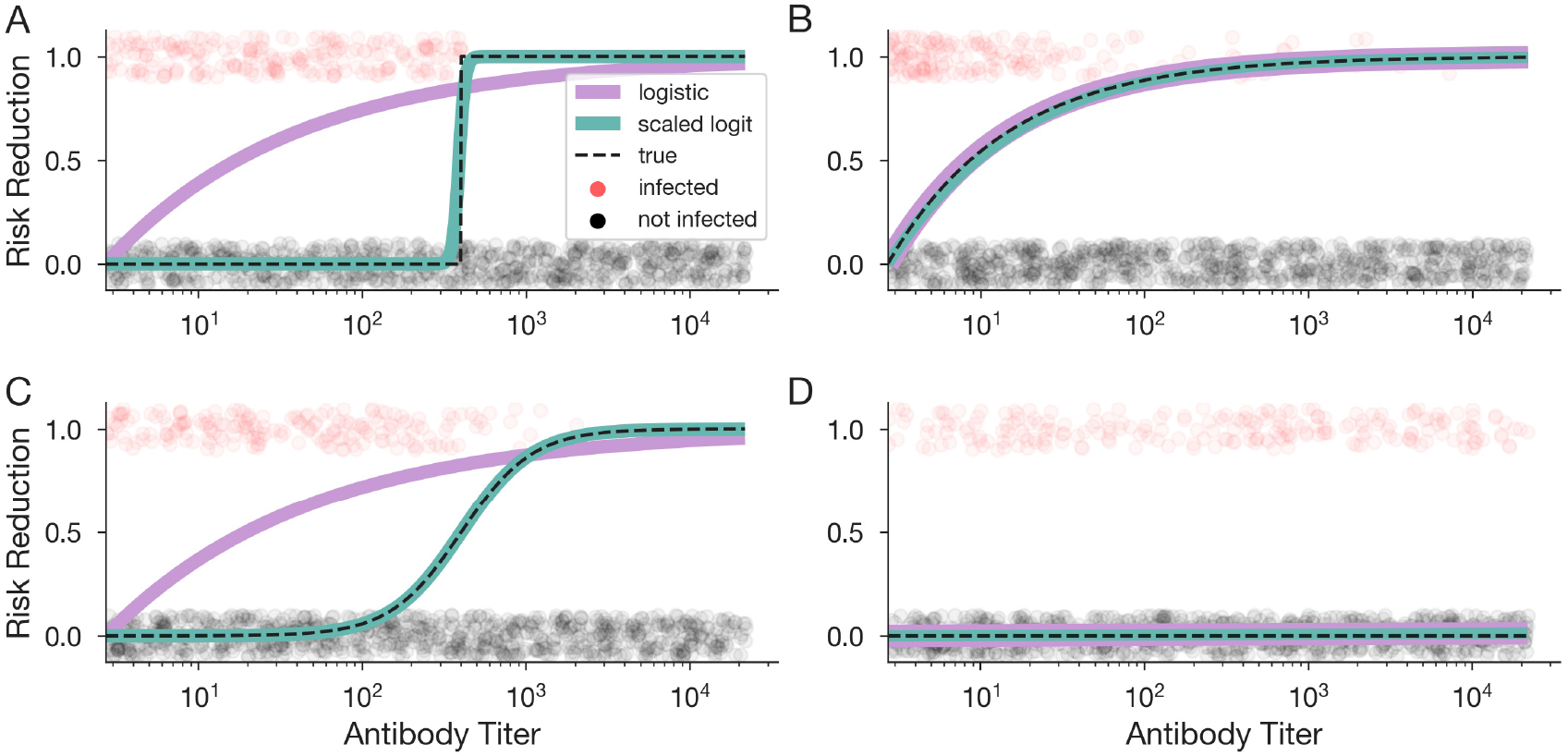
The scaled logit model can recover more general antibody protection functions, while logistic regression is limited in its scope. Estimated protection function using the scaled logit model (green curve) trained on data generated using a threshold (A), exponential (B), sigmoidal (C), and flat (D) protection functions (dashed black curve). Circles show a subset of infected (red) and uninfected (gray) simulated individuals at various antibody titers used to train the model.

**Figure S2:**
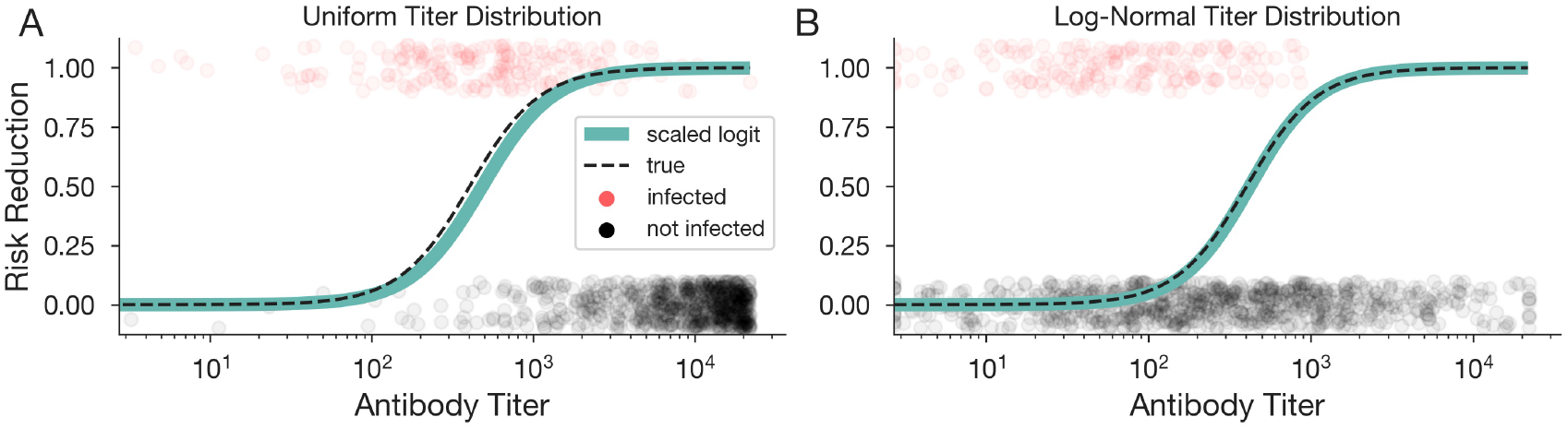
The scaled logit model can recover more general antibody protection functions, even under different titer distributions. Estimated protection function using the scaled logit model (green curve) trained on data generated using exponential (A) and sigmoidal (B) protection functions (dashed black curve) for uniformly (A) and normally (B) distributed titers. Circles show a subset of infected (red) and uninfected (gray) simulated individuals at various antibody titers used to train the model.

**Figure S3:**
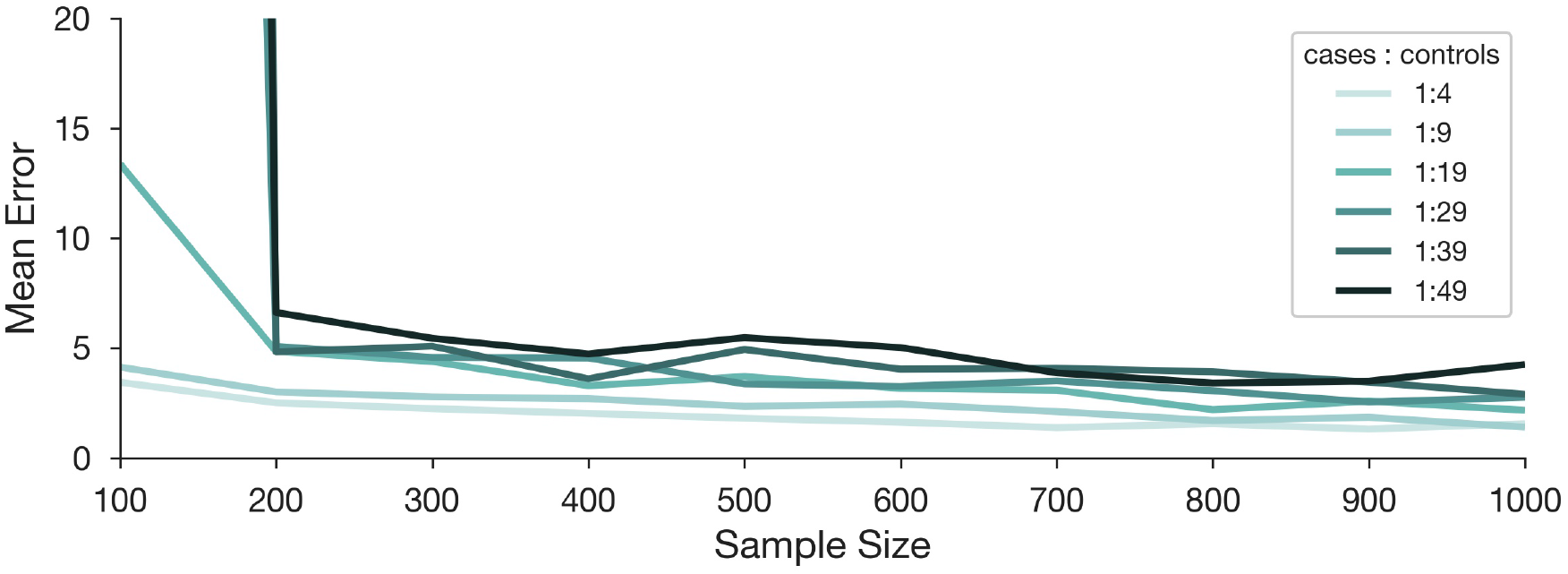
The scaled logit model has low accuracy at small sample sizes. Average error over 50 simulations per scenario for various sample sizes and case-to-control ratios using the scaled logit model. Error was defined as the discrete *ℓ*_2_ norm (Euclidean distance) between the modeled and true protection functions. Darker-colored lines represent scenarios with more controls per case.

**Figure S4:**
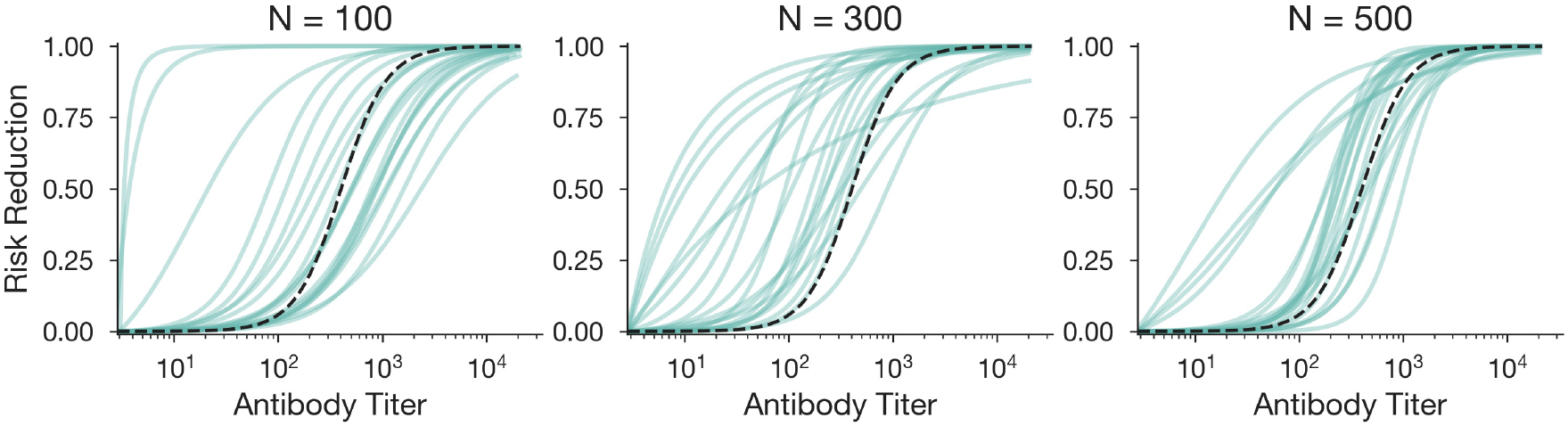
Scaled logit model fit with low sample sizes. Estimated protection function using the scaled logit model (green curves) for 50 stochastic simulations at the specified sample size (*N*) using a sigmoidal true protection (dashed black curve).

**Figure S5:**
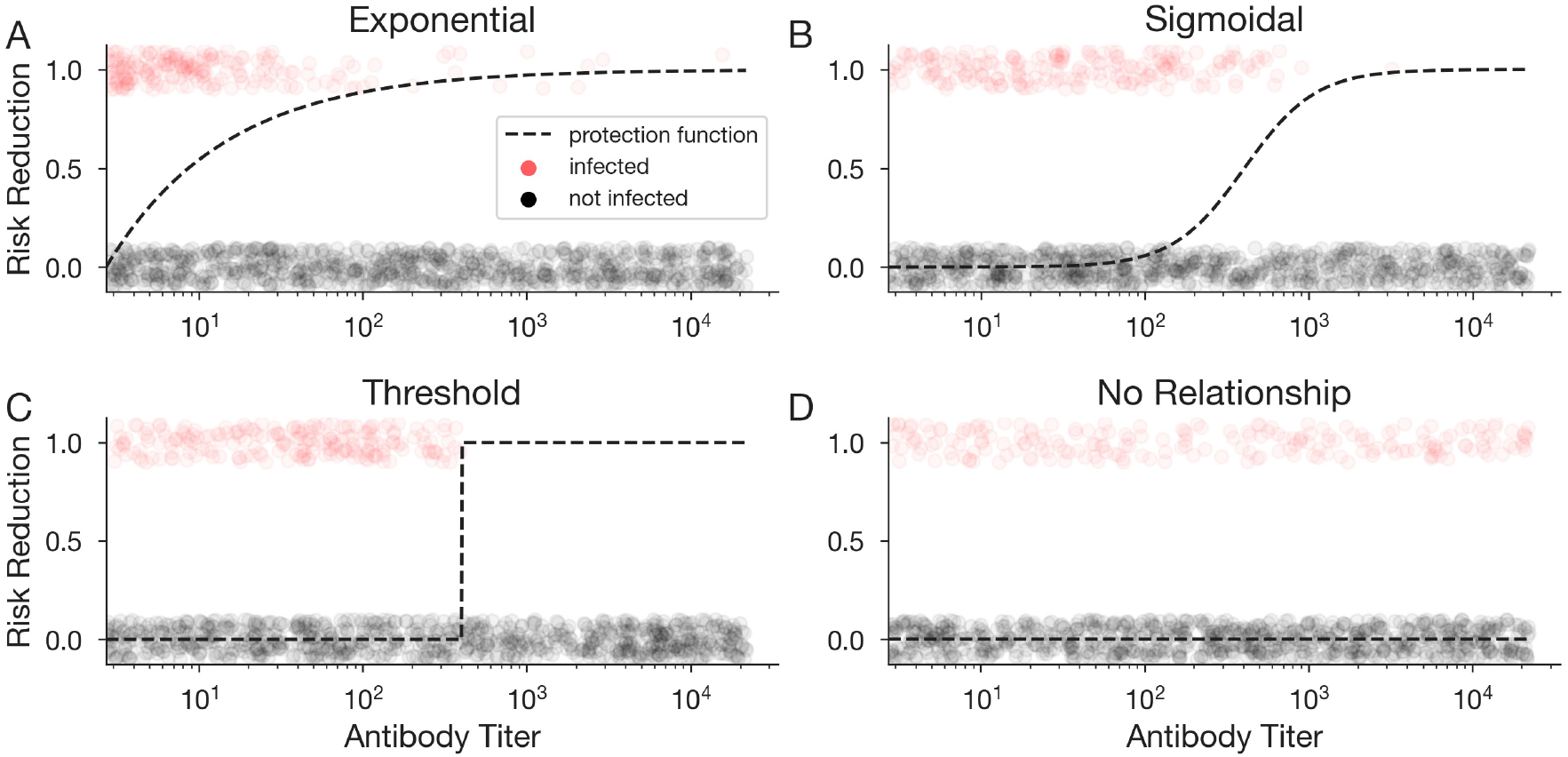
Protection functions used for data simulation. Risk reduction as a function of antibody titer for exponential (A), sigmoidal (B), threshold (C), and no relationship (D) protection functions.

## Fitting the scaled logit model to data

We use the built in `scipy.minimize` function in python, implemented with the Nelder-Mead minimization algorithm [35] and options to increase the number of iterations before convergence. In python, this process uses the following code, where *λ* has been replaced with *k* to avoid conflict with built-in functions:

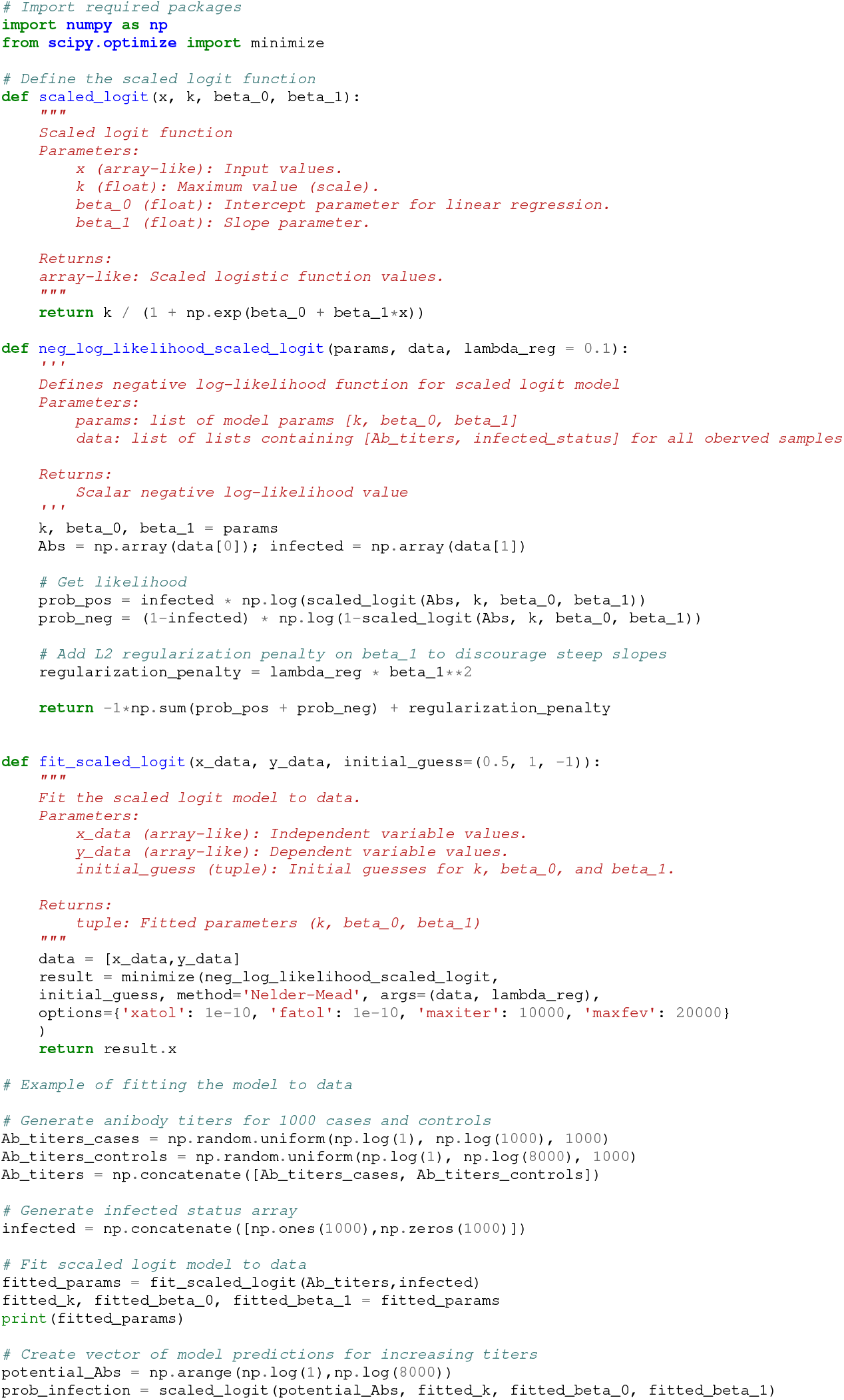

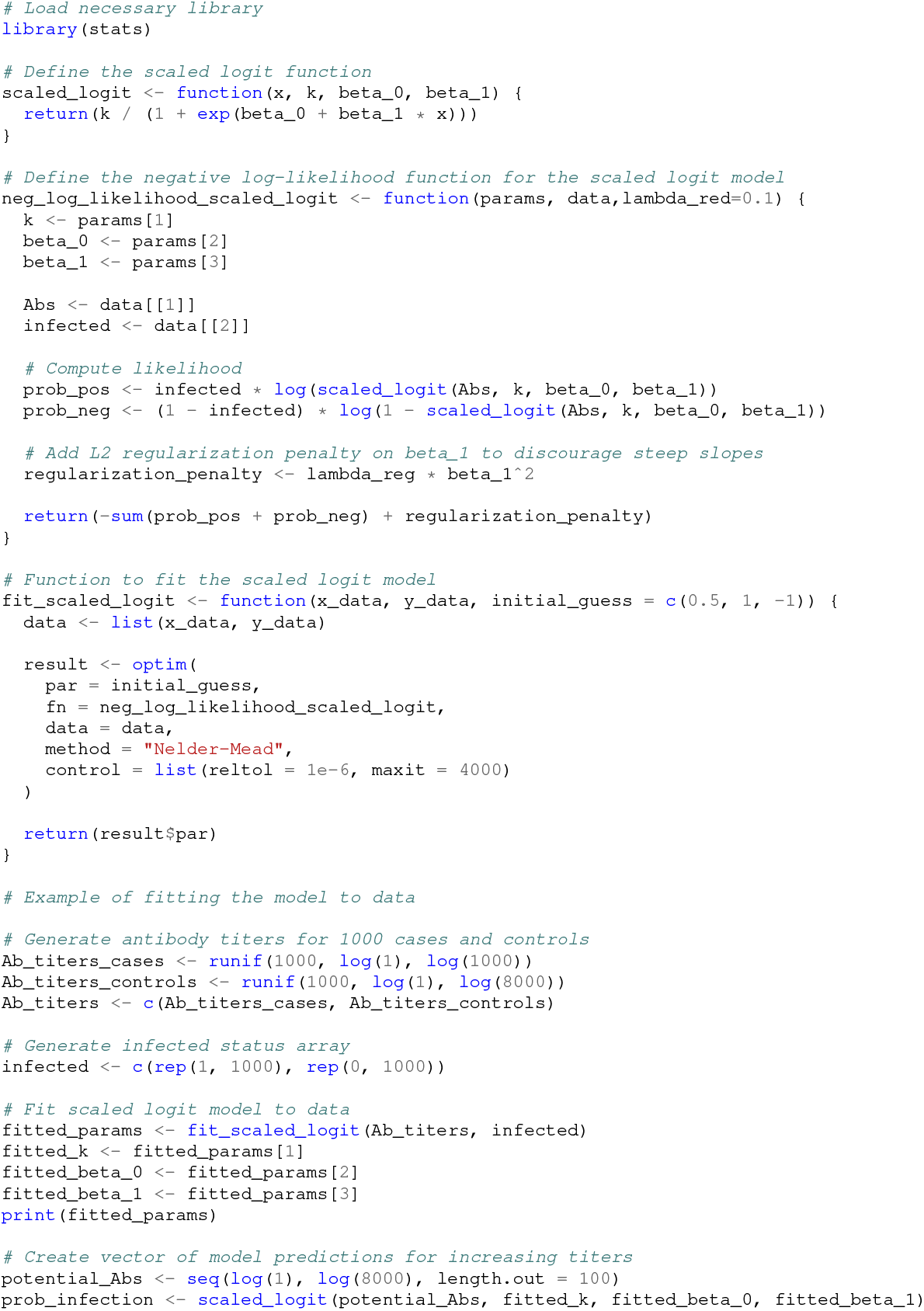

## References

[1] Arnold S. Monto, Joshua G. Petrie, Rachel T. Cross, Emileigh Johnson, Merry Liu, Weimin Zhong, Min Levine, Jacqueline M. Katz, and Suzanne E. Ohmit. Antibody to influenza virus neuraminidase: An independent correlate of protection. The Journal of Infectious Diseases, 212(8):1191–1199, 2015.

[2] Eric J. Nilles, Cecilia Then Paulino, Michael de St Aubin, William Duke, Petr Jarolim, Isaac Miguel Sanchez, Kristy O Murray, Colleen L Lau, Emily Zielinski Gutiérrez, Ronald Skewes Ramm, Marietta Vasquez, and Adam Kucharski. Tracking immune correlates of protection for emerging SARS-CoV-2 variants. Lancet Infectious Diseases, 2023.

[3] Li Qin, Peter B. Gilbert, Lawrence Corey, M. Juliana McElrath, and Steven G. Self. A framework for assessing immunological correlates of protection in vaccine trials. The Journal of Infectious Diseases, 196(9):1304–1312, 2007.

[4] Alexandra B. Hogan, Patrick Doohan, Sean L. Wu, Daniela Olivera Mesa, Jaspreet Toor, Oliver J. Watson, Peter Winskill, Giovanni Charles, Gregory Barnsley, Eleanor M. Riley, David S. Khoury, Neil M. Ferguson, and Azra C. Ghani. Estimating long-term vaccine effectiveness against SARS-CoV-2 variants: a model-based approach. Nature Communications, 14(1):4325, 2023.

[5] Michal Canetti, Noam Barda, Yaniv Lustig, Yael Weiss-Ottolenghi, Victoria Indenbaum, Yovel Peretz, Neta Zuckerman, Keren Asraf, Sharon Amit, Yitshak Kreiss, and Gili Regev-Yochay. Risk factors and correlates of protection against XBB SARS-CoV-2 infection among health care workers. Vaccine, 42(26):126308, 2024.

[6] Can Liu, Jiawei Zhang, Yongbin Zeng, Chun Huang, Falin Chen, Yingping Cao, Siying Wu, Donghong Wei, Zhong Lin, Yali Zhang, Ling Zhang, Jing Teng, Zishun Li, Guolin Hong, Tianci Yang, Huiming Ye, Haijian Tu, Yupeng Xiao, Lishan Huang, Caorui Lin, Tianbin Chen, Yanqin Deng, Qishui Ou, and Jinming Li. Effectiveness of SARS-CoV-2-inactivated vaccine and the correlation to neutralizing antibodies: A test-negative case–control study. Journal of Medical Virology, 95(1):e28280, 2023.

[7] David S. Khoury, Deborah Cromer, Arnold Reynaldi, Timothy E. Schlub, Adam K. Wheatley, Jennifer A. Juno, Kanta Subbarao, Stephen J. Kent, James A. Triccas, and Miles P. Davenport. Neutralizing antibody levels are highly predictive of immune protection from symptomatic SARS-CoV-2 infection. Nature Medicine, 27(7):1205–1211, 2021.

[8] Erica Ollmann Saphire, Sharon L. Schendel, Bronwyn M. Gunn, Jacob C. Milligan, and Galit Alter. Antibody-mediated protection against Ebola virus. Nature Immunology, 19(11):1169–1178, 2018.

[9] Julie A. Wilson, Michael Hevey, Russell Bakken, Shawn Guest, Mike Bray, Alan L. Schmaljohn, and Mary Kate Hart. Epitopes involved in antibody-mediated protection from Ebola virus. Science, 287(5458):1664–1666, 2000.

[10] John R Mascola. Passive transfer studies to elucidate the role of antibody-mediated protection against HIV-1. Vaccine, 20(15):1922–1925, 2002.

[11] Joseph A Lewnard, Christine Tedijanto, Benjamin J Cowling, and Marc Lipsitch. Measurement of vaccine direct effects under the test-negative design. American Journal of Epidemiology, 187(12):2686–2697, 2018.

[12] Michael L. Jackson and Jennifer C. Nelson. The test-negative design for estimating influenza vaccine effectiveness. Vaccine, 31(17):2165–2168, 2013.

[13] Natalie E. Dean, Joseph W. Hogan, and Mireille E. Schnitzer. Covid-19 vaccine effectiveness and the test-negative design. New England Journal of Medicine, 385(15):1431–1433, 2021.

[14] Wakaba Fukushima and Yoshio Hirota. Basic principles of test-negative design in evaluating influenza vaccine effectiveness. Vaccine, 35(36):4796–4800, 2017.

[15] Erik K Johnson, Rebecca Kahn, Yonatan H Grad, Marc Lipsitch, and Daniel B Larremore. Test negative designs with uncertainty, sensitivity, and specificity. medRxiv, 2021.

[16] Sheena G. Sullivan, Eric J. Tchetgen, and Benjamin J. Cowling. Theoretical basis of the test-negative study design for assessment of influenza vaccine effectiveness. American Journal of Epidemiology, 184(5):345–353, 2016.

[17] George R. Siber, Ih Chang, Sherryl Baker, Philip Fernsten, Katherine L. O’Brien, Mathuram Santosham, Keith P. Klugman, Shabir A. Madhi, Peter Paradiso, and Robert Kohberger. Estimating the protective concentration of anti-pneumococcal capsular polysaccharide antibodies. Vaccine, 25(19):3816–3826, 2007.

[18] Ana P. Goncalvez, Cheng-Hsin Chien, Kamolchanok Tubthong, Inna Gorshkova, Carrie Roll, Olivia Donau, Peter Schuck, Sutee Yoksan, Sy-Dar Wang, Robert H. Purcell, and Ching-Juh Lai. Humanized monoclonal antibodies derived from chimpanzee fabs protect against japanese encephalitis virus in vitro and in vivo. Journal of Virology, 82(14):7009–7021, 2008.

[19] David S. Khoury, Timothy E. Schlub, Deborah Cromer, Megan Steain, Youyi Fong, Peter B. Gilbert, Kanta Subbarao, James A. Triccas, Stephen J. Kent,, and Miles P. Davenport. Correlates of protection, thresholds of protection, and immunobridging among persons with SARS-CoV-2 infection. Emerging Infectious Diseases journal - CDC, 29(2), 2023.

[20] Andrew J. Dunning. A model for immunological correlates of protection. Statistics in Medicine, 25(9):1485–1497, 2006.

[21] Sheena G. Sullivan and Benjamin J. Cowling. “crude vaccine effectiveness” is a misleading term in test-negative studies of influenza vaccine effectiveness. Epidemiology, 26(5):e60, 2015.

[22] Ziyuan Zhang, Christopher Brian Boyer, and Marc Lipsitch. Use of the test-negative design to estimate the protective effect of a scalar immune measure: A simulation analysis. medRxiv, 2024.

[23] Eric J Nilles, Cecilia Then Paulino, Michael de St Aubin, William Duke, Petr Jarolim, Isaac Miguel Sanchez, Kristy O Murray, Colleen L Lau, Emily Zielinski Gutiérrez, Ronald Skewes Ramm, Marietta Vasquez, and Adam Kucharski. Tracking immune correlates of protection for emerging SARS-CoV-2 variants. The Lancet Infectious Diseases, 23(2):153–154, 2023.

[24] Andrew J. Dunning, Jennifer Kensler, Laurent Coudeville, and Fabrice Bailleux. Some extensions in continuous models for immunological correlates of protection. BMC Medical Research Methodology, 15(1):107, 2015.

[25] Akira Endo, Sebastian Funk, and Adam J Kucharski. Bias correction methods for test-negative designs in the presence of misclassification. Epidemiology & Infection, 148:e216, 2020.

[26] Alexander Muik, Bonny Gaby Lui, Jasmin Quandt, Huitian Diao, Yunguan Fu, Maren Bacher, Jessica Gordon, Aras Toker, Jessica Grosser, Orkun Ozhelvaci, Katharina Grikscheit, Sebastian Hoehl, Niko Kohmer, Yaniv Lustig, Gili Regev-Yochay, Sandra Ciesek, Karim Beguir, Asaf Poran, Isabel Vogler, özlem Türeci, and Ugur Sahin. Progressive loss of conserved spike protein neutralizing antibody sites in omicron sublineages is balanced by preserved T cell immunity. Cell Reports, 42(8), 2023.

[27] Avni Vatti, Diego M. Monsalve, Yair Pacheco, Chia Chang, Juan-Manuel Anaya, and M. Eric Gershwin. Original antigenic sin: A comprehensive review. Journal of Autoimmunity, 83:12–21, 2017.

[28] Scott B Halstead. Neutralization and antibody-dependent enhancement of dengue viruses. Adv Virus Res., 60:421–67, 2003.

[29] Daniel Westreich and Michael G. Hudgens. Invited commentary: Beware the test-negative design. American Journal of Epidemiology, 184(5):354–356, 2016.

[30] Marcus Carlsson, Jens Wittsten, and Cecilia Söderberg-Nauclér. A note on variable susceptibility, the herd-immunity threshold and modeling of infectious diseases. PLOS ONE, 18(2):e0279454, 2023.

[31] Joel C. Miller. Epidemic size and probability in populations with heterogeneous infectivity and susceptibility. Physical Review E, 76(1):010101, 2007.

[32] Antonio Montalbán, Rodrigo M. Corder, and M. Gabriela M. Gomes. Herd immunity under individual variation and reinfection. Journal of Mathematical Biology, 85(2), 2022.

[33] Eva Kisdi. Optimal vaccination strategies for imperfect vaccines and variable host susceptibility. Journal of Theoretical Biology, 594:111899, 2024.

[34] Sophie L. Larsen, Junke Yang, Huibin Lv, Yang Wei Huan, Qiwen Teo, Tossapol Pholcharee, Ruipeng Lei, Akshita B Gopal, Evan K. Shao, Logan Talmage, Chris K. P. Mok, Saki Takahashi, Alicia N. M. Kraay, Nicholas C. Wu, and Pamela P. Martinez. Reimagining the serocatalytic model for infectious diseases: a case study of common coronaviruses. medRxiv, 2024.

[35] Fuchang Gao and Lixing Han. Implementing the Nelder-Mead simplex algorithm with adaptive parameters. Computational Optimization and Applications, 51(1):259–277, 2012.

